# Donor Anti-Spike Immunity is Related to Recipient Recovery and Can Predict the Efficacy of Convalescent Plasma Units

**DOI:** 10.1101/2021.02.25.21252463

**Authors:** Sanath Kumar Janaka, William Hartman, Huihui Mou, Michael Farzan, Susan L. Stramer, Erin Goodhue, John Weiss, David Evans, Joseph P. Connor

## Abstract

**Background:** The novel coronavirus, SARS-CoV2 that causes COVID-19 has resulted in the death of more than 2.31 million people within the last year and yet no cure exists. Whereas passive immunization with COVID-19 convalescent plasma (CCP) provides a safe and viable option, selection of optimal units for therapy and lack of clear therapeutic benefit from transfusion remain as barriers to the use of CCP.

**Study design and methods:** To identify plasma that is expected to benefit recipients, we measured anti-SARS-CoV2 antibody levels using clinically available serological assays and correlated with the neutralizing activity of CCP from donors. Neutralizing titer of plasma samples was measured by assaying infectivity of SARS-CoV-2 spike protein pseudotyped retrovirus particles in the presence of dilutions of plasma samples. We also used this assay to identify evidence of passive transfusion of neutralizing activity in CCP recipients.

**Results:** Viral neutralization and anti-spike protein antibodies in 109 samples from 87 plasma donors were highly varied but modestly correlated with each other. Recipients who died of COVID-19 were found to have been transfused with units with lower anti-spike antibody levels and neutralizing activity. Passive transfer of neutralization activity was documented in 62% of antibody naive plasma recipients.

**Conclusions:** Since viral neutralization is the goal of CCP transfusion, our observations not only support the use of anti-spike SARS-CoV2 serology tests to identify beneficial CCP units, but also support the therapeutic value of convalescent plasma with high titers of anti-spike antibodies.

## Introduction

Since its emergence in December 2019, SARS-CoV2, the causative agent of COVID-19 has infected over 108 million people and has been responsible for the death of more than 2.3 million persons globally with over 27 million infections and greater than 485,000 deaths in the United States (Johns Hopkins Coronavirus Resource Center, online Nov. 2020). At this time no curative therapies are available thus passive immunization with COVID-19 convalescent plasma (CCP) remains a viable therapeutic option. Despite preliminary evidence of both safety and clinical efficacy ^1-6^, the antibody response in terms of titer, viral target, and neutralization activity required for the therapeutic effect of CCP remains incomplete ^7-9^ leaving the identification of optimal CCP units for transfusion undefined.

In order to better define the immune response to SARS-CoV2 and how this may be used to optimize the role of CCP we studied the activity of SARS-CoV2 neutralizing antibodies in samples from CCP donors using a retrovirus-pseudo-type neutralization assay, compared these results with antibody levels from two clinically used serologic antibody tests, explored whether passive neutralization activity can be identified in patients after transfusion of CCP, and explored the relationship between these immunologic assessments in donor units and clinical outcomes in CVOID-19 patients who were transfused CCP.

## Materials and Methods

### Cell lines

293T cells were obtained from ATCC and maintained in DMEM with 10% FBS, 100 U/ml penicillin, 100 µg/ml streptomycin, 0.25 µg/ml amphotericin B, and 2mM L-glutamine (D10). Plasmid expressing the human ACE2 protein with a C-terminal C9 tag was obtained from Addgene (Plasmid 1786) ^12^. 293T cells were then transduced with retroviral particles carrying a pQCXIP vector cloned to encode human ACE2 protein, selected and maintained in D10 supplemented with 1µg/ml Puromycin. ACE2 expression was confirmed by infection of 293T-ACE2 cells with retroviruses and lentiviruses pseudotyped with SARS-CoV-1 S and SARS-CoV2 S proteins.

### Neutralization Assay

The neutralization assay was performed as described previously using plasmids generously provided by Michael Farzan ((Scripps Research Institute, FL) ^10,11^.SARS-CoV-2 spike protein pseudotyped MLV particles carrying a firefly Luciferase reporter as the infectious agent. The virus stocks were titrated on 293T-ACE2 cells to determine an optimal volume of the virus to be used in neutralization assays. Plasma and serum samples were obtained from the American Red Cross or the UW Carbone Cancer Center. EDTA anticoagulant was removed from plasma samples by dialysis against phosphate buffered saline (PBS) using 10MWCO Slide-A-lyzer dialysis cassettes. Infectivity was measured relative to the untreated wells after subtracting the signal from uninfected wells. Each instance of the assay run included plasma from known COVID-19 negative donors as a negative control. The dilution of sample resulting in 50% infectivity was determined and reported as the IC50 neutralizing titer.

### Anti-SARS-CoV2 antibody/serology assays

Anti-SARS-CoV2 Spike antibodies (Ortho Diagnostics, Markham, Ontario): This assay is a qualitative, automated, two stage immunometric assay to identify both anti-SARS-CoV2 Spike protein IgG and IgM. The assay provides a signal to cut-off value (S/CO) with a manufacturer defined threshold value to indicate a reactive sample.

Architect Anti-SARS-CoV2 nucleocapsid IgG assay (Abbott Laboratories, Chicago, IL): This is an automated, two-step immunoassay for the detection of IgG antibodies to SARS-CoV2 nucleocapsid proteins. The presence of IgG antibodies are quantitated as a signal to cut off INDEX (Abbott INDEX) with a manufacturer defined threshold defining a reactive sample. Although neither of these assays are used clinically as quantitative tests for the purposes of this study both the S/CO and the INDEX were considered to define the amount of anti-SARS-CoV2 antibodies in each sample.

CCP recipients and clinical parameters: This study was approved by the University of Wisconsin Institutional Review Board. All cases met criteria for enrollment in the Mayo Clinic Expanded Access Protocol (IND # 20-003312, EAP) and gave written, informed consent for CCP transfusion and data collection. All patients had laboratory-confirmed COVID-19 by RT-PCR with either severe or life-threatening disease as described previously ^2-4^.

CCP was collected in collaboration with the American Red Cross (ARC) and serum and plasma samples were collected at the time of donation, aliquoted and stored frozen for analysis. Residual samples from clinical laboratory testing in hospitalized COVID-19 patients were collected, aliquoted and stored frozen by the staff of the University of Wisconsin’s Comprehensive Cancer Center Biobank under a previously approved IRB approved protocol. Recipient data were abstracted from the electronic medical record. Time to disease escalation was defined as the time, in days, from admission to the date when there was sustained need (12-24 hours) for an increased level of oxygen/respiratory support. Similarly, the time to respiratory improvement was defined as the time from of admission or transfusion, in days, to the date when the patient demonstrated a sustained (12-24 hours) decrease in oxygen requirements.

### Statistical Analysis

Data from plasma and serum samples were combined together after validation of neutralization assays. Results were presented with descriptive statistics and parametric and non-parametric tests as appropriate. All statistical analysis was done using the GraphPad Prism software (GraphPad Software, Inc La Jolla, CA).

## Results

One-hundred and-nine samples from 87 individual COVID-19 convalescent plasma donors were studied including 56 serum samples and 53 plasma samples. As demonstrated by the wide standard deviations (SDs) in table 1 the neutralization activity defined by IC50 values varied widely in plasma units with titers ranging from < 8 to 581. Similarly, both the Ortho total Ig S/CO values and the Abbott INDEX value varied widely with ranges from 0.02 to 904.0 and 0.9 to 8.7 respectively. None of the three immune parameters were found to be associated with the general donor characteristics of age, gender, or ABO/Rh blood grouping (ANOVA, data not shown)

**Table 1.** General Characteristics and Immune Response Parameters of 87 CCP Donors

|  |  |
| --- | --- |
| Gender (N, (% total group)) | Male 39 (45%)<br>Female 48 (55%) |
| Age (mean ± SD) | 47.2 ± 14.9 years<br>(median 47, range 20-80 years) |
| ABO Group (N, (% total group)) | A 32 (37%, 78%Rh +)<br>B 16 (18%, 69% Rh +)<br>AB 3 (3%, 100% Rh +)<br>O 36 (42%, 92% Rh +) |
| Neutralization IC50 (mean ± SD) | 121.1 ± 133.9<br>(median 61.4, range <8.0-581.4)<br>Quartile 1 <8 – 31<br>Quartile 2 32 – 61<br>Quartile 3 62 – 153<br>Quartile 4 154-581 |
| Ortho (Ig total) S/CO value (mean ± SD) | 229.6 ± 188.4<br>(median 203, range 0.02-904)<br>Quartile 1 0.02 – 49.8<br>Quartile 2 50 – 203<br>Quartile 3 204 – 352<br>Quartile 4 353 - 904 |
| Abbott (IgG) INDEX value (mean ± SD) | 4.8 ± 2.0<br>(median 4.6, range 0.9-8.7)<br>Quartile 1 0.9 – 2.9<br>Quartile 2 3.0 – 4.6<br>Quartile 3 4.7 – 6.8<br>Quartile 4 6.9 – 8.7 |

Twenty-two donors had both a serum and a plasma sample for independent neutralization testing and the neutralization assay was validated by comparing the IC50 values of matched plasma and serum samples. IC50 values for the two different sample types were strongly correlated (r = 0.86 <0.0001). Consistent IC50 values with different samples from the same donor both validates our neutralization assay and provides a rationale to combine results from both serum and plasma samples in our analysis.

Both the Ortho S/CO value and the Abbott INDEX modestly correlate with the IC50 for each donor unit with r= 0.37 and r = 0.40 respectively. Similarly, the Ortho S/CO and the Abbott INDEX modestly correlate with one another, r = 0.33.

To extend the study further, neutralizing responses in plasma transfused to COVID-19 recipients were analyzed. From April to August 2020, 48 hospitalized patients were transfused CCP at the University of Wisconsin Hospital. One pediatric patient, one patient still acutely hospitalized at the time of analysis, and two other patients incidentally found to be COVID-19 positive were excluded as they were admitted with a non-COVID-19, life-threatening illness (one acute leukemia and one decompensated valvular heart disease). The general characteristics of the study group’s hospitalization and the course of their respiratory disease is summarized in table 2. Most (80%) of the patients were admitted from home with an average duration of symptoms prior to admission of 8.3 days and 80% were classified as having severe disease on admission.

**Table 2.** Clinical Demographics and Hospital Course for Patients Transfused CCP.

| General Clinical Demographics |  |
| --- | --- |
| Age (mean ± SD) | 60.2 ± 13.7 years<br>(median 61, range 22-89 years) |
| Gender (N (% total group)) | Male 22 (50%) Female 22 (50%) |
| ABO Blood Type<br>(N (% total group)) | A 16 (36%)<br>B 1 (2%)<br>AB 2 (5%)<br>O 25 (57%) |
| Rh (N (% total group)) | Positive 36 (82%) Negative 8 (18%) |
| Admission and Hospitalization Details |  |
| Residence Prior to Admission | Home (self-care) 35 (80%)<br>Skilled Nursing Facility 9 (20%) |
| Duration of Symptoms Prior to Admission<br>(mean ± SD) | 8.3 ± 5 days<br>(median 7, range 2-28 days) |
| COVID-19 Disease Status on Admission | Severe Disease 36 (82%)<br>Life-Threatening Disease 8 (18%) |
| Hospital Course of COVCID-19 |  |
| Escalation of Oxygen Requirements | YES 16 (36%)<br>NO 18 (41%)<br>Escalation on admit 10 (23%)<br>(Escalation within 24 hours of Admission) |
| Admission to Escalation in days (mean ± SD) | 3.9 ± 3.0 days<br>(median 3.0, range 1-10 days) |
| Intubation/Mechanical Ventilation | YES 17 (39%)<br>NO 27 (61%) |
| Duration of Mechanical Ventilation in days<br>(mean ± SD, excludes 5 deaths while ventilated) | 10.5 ± 8.4 days<br>(median 7, range 1-24 days) |
| Admission to Respiratory Improvement in days<br>(mean ± SD) | 7.9 ± 5.4 days<br>(median 6.0, range 1-23 days) |
| Other Treatments rendered | Hydroxychloroquine 10 (23%)<br>Systemic Steroids 22 (50%)<br>Remdesivir 22 (50%)<br>Antibiotics 27 (67%)<br>Vassopressors 14 (32%) |
| Outcome/Discharge Details |  |
| Length of Stay (mean ± SD) | 15.8 ± 11.3 days<br>(median 10.5, range 3-40 days) |
| Discharge Status | Home Self-care 23 (52%)<br>Skilled Nursing Facility 11 (25%)<br>Rehab. Facility 3 (7%)<br>DIED (in-hospital death) 7 (16%) |
| Oxygen Requirements at Discharge | None 28 (64%)<br>NC O2 7 (16%)<br>Died 7 (16%) |

Seventeen patients (39%) required intubation and mechanical ventilation during their hospitalization. Eight of these patients were intubated on admission or were transferred intubated from an outside hospital. Six cases required intubation within the first 24 hours of admission and one patient was intubated 3 days after admission just prior to convalescent plasma. All of these 15 cases were transfused CCP after intubation. Two patients (5%) progressed to requiring mechanical ventilation after transfusion of CCP, one at 4 days and the other at 7 days post-transfusion. The average duration of mechanical ventilation was 10.5 days. Exclusive of those that died during their hospitalization, most showed respiratory improvement at a mean of 7.9 days from admission. All patients were treated with at least one additional treatment for COVID-19 including Hydroxychloroquine, systemic steroids and anti-retroviral drugs. The average length of stay was 15.8 days.

Seven subjects (16%) had died of COVID after convalescent plasma transfusion and 37 patients (84%) were discharged alive.

Of the 48 transfused patients a sample from the donor unit/units transfused was available for immunologic testing in 44 (92%) cases. The transfusion details for the final 44 cases studied are summarized in table 3. As was seen for the entire donor population studied (table 1), the serologic testing and neutralization IC50 varied widely among the units transfused to patients.

**Table 3.** COVID-19 Convalescent Plasma Transfusion Details.

|  |  |
| --- | --- |
| Admitted before CCP units were available | YES 9 (20%)<br>NO 35 (80%) |
| Admission to Transfusion in days (mean + SD) | Admit Prior to CCP available: $9.0 \pm 6.7$ days<br>(median 9.0, range 2-22 days)<br>Admit after CCP available: $2.1 \pm 2.1$ days<br>(median 1.0, range 0-10 days) |
| Number of Units Transfused<br>(2 units for weight > 90Kg) | 1 Unit 36 (82%)<br>2 Units 8 (18%) |
| Transfusion to Respiratory Improvement in days<br>(mean ± SD) | $4.4 \pm 4.2$ days<br>(median 3.0, range -3-16 days) |
| Transfusion to Extubation, N = 12<br>(mean + SD, excludes 5 deaths while ventilated) | $5.8 \pm 6.2$ days<br>(median 3.0, range -3-10 days) |
| Neutralization IC50 (mean ± SD) | $177.5 + 176.1$<br>(median 106.4, range 21.7-559.5) |
| Ortho (Ig total) S/CO value (mean ± SD) | $135.8 + 136.6$<br>(median 60.7, range 1.2-442.0) |
| Abbott (IgG) INDEX value (mean ± SD) | $5.2 + 1.9$<br>(median 5.2, range 1.8-8.7) |

Early in the study patients were admitted to the hospital prior to the availability of convalescent plasma units for transfusion. Subsequently these patients (N=9) were transfused relatively late at a mean of 9 days after admission. After routine availability most transfusions (N=35) were provided within 2 days of admission (table 3). Although the mortality rate was lower in those transfused earlier in their hospitalization, 14% vs 22%, this did not achieve statistical significance (p =0.25 by Chi Square). Furthermore, no significant associations were found between the immune response results (as two groups around the median or as quartiles) and the time to either clinical improvement or hospital discharge (by t-test or ANOVA, data not shown).

To study the benefit of CCP transfusion, we compared the antibody levels and donor neutralization activity transfused to recipients at the extremes of COVID-19 outcomes, those who died in the hospital (N = 7) and those discharged home without the need for ongoing oxygen support (N = 18). Mean IC50 and anti-spike antibody levels were lower in patients who died versus those who were discharged, IC50 of 63.2 ± 45.4 vs 152.4 ± 161.3 (p = 0.05 figure 1A), and S/CO value of 85.5 ± 84 vs 223.2 ± 132 (figure 1B, p = 0.018). In contrast, the Abbott IgG INDEX showed no significant differences between the two groups 4.97 ± 1.3 vs 5.02 ± 2.0 (figure 1C, p = 0.95). Further evaluation of the units transfused to patients who died showed that four out of seven units had Ortho Ig total S/CO values within the lowest quartile based on the entire donor population (table 1).

**Figure 1.**
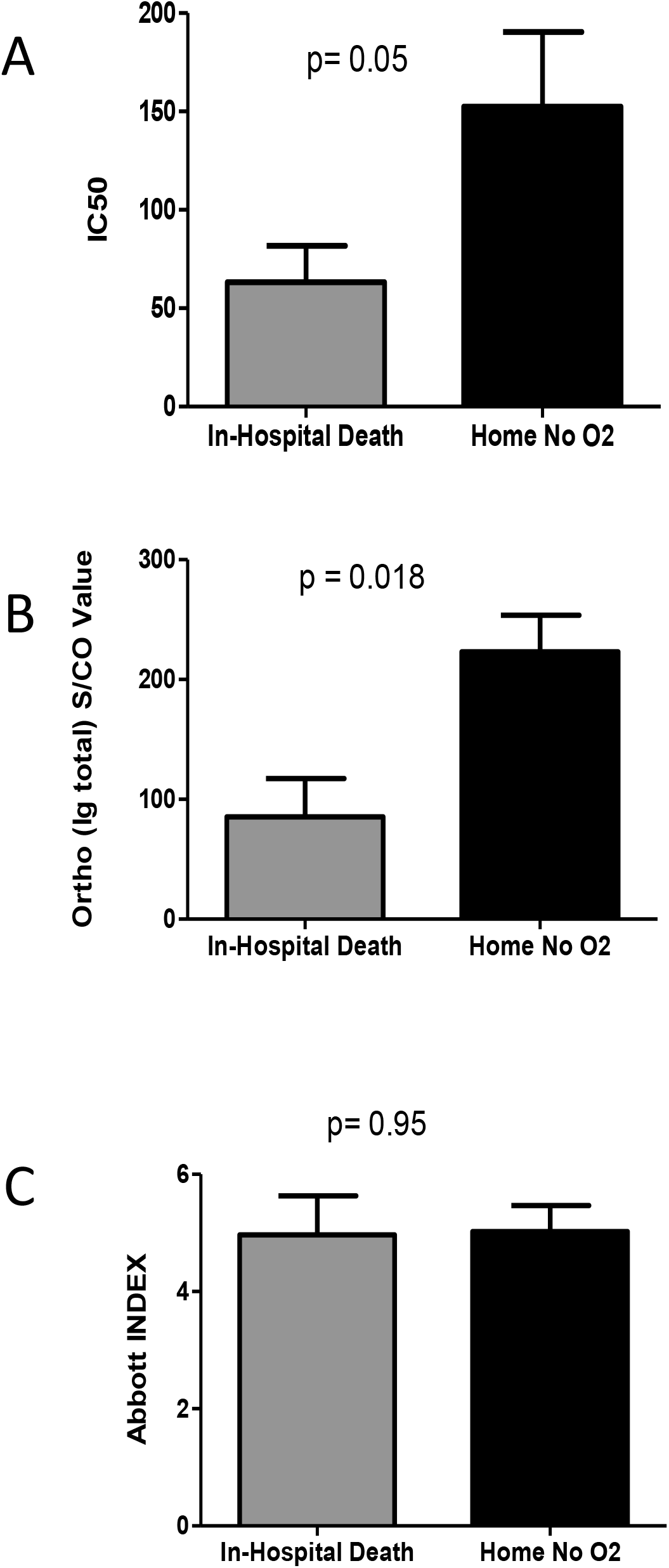
COVID-19 outcome-based comparison of neutralization activity and antibody responses in transfused plasma units. Mean values of neutralization IC50 (A), anti-spike total Ig S/CO values (B), and anti-nucleoprotein IgG INDEX values (C) of transfused units were compared among plasma recipients at the extremes of COVID-19 outcomes, death in hospital or discharge without oxygen requirement. *p*-values were compared using Mann-Whitney tests.

To further explore the relationship between the Ortho, anti-spike protein serology and neutralization IC50, each outcome variable was analyzed against the other in quartiles for all 87 plasma donor cases. This analysis shows that for optimal therapeutic effect of CCP, the first quartile and at least half of the second quartile samples with Ortho S/CO value range (table 1) should be excluded (figure 2A) and that this corresponds to the exclusion of the entire lowest quartile based on neutralization activity (figure 2B). The Abbott INDEX value did not show any difference among quartiles based on IC50 (data not shown).

**Figure 2.**
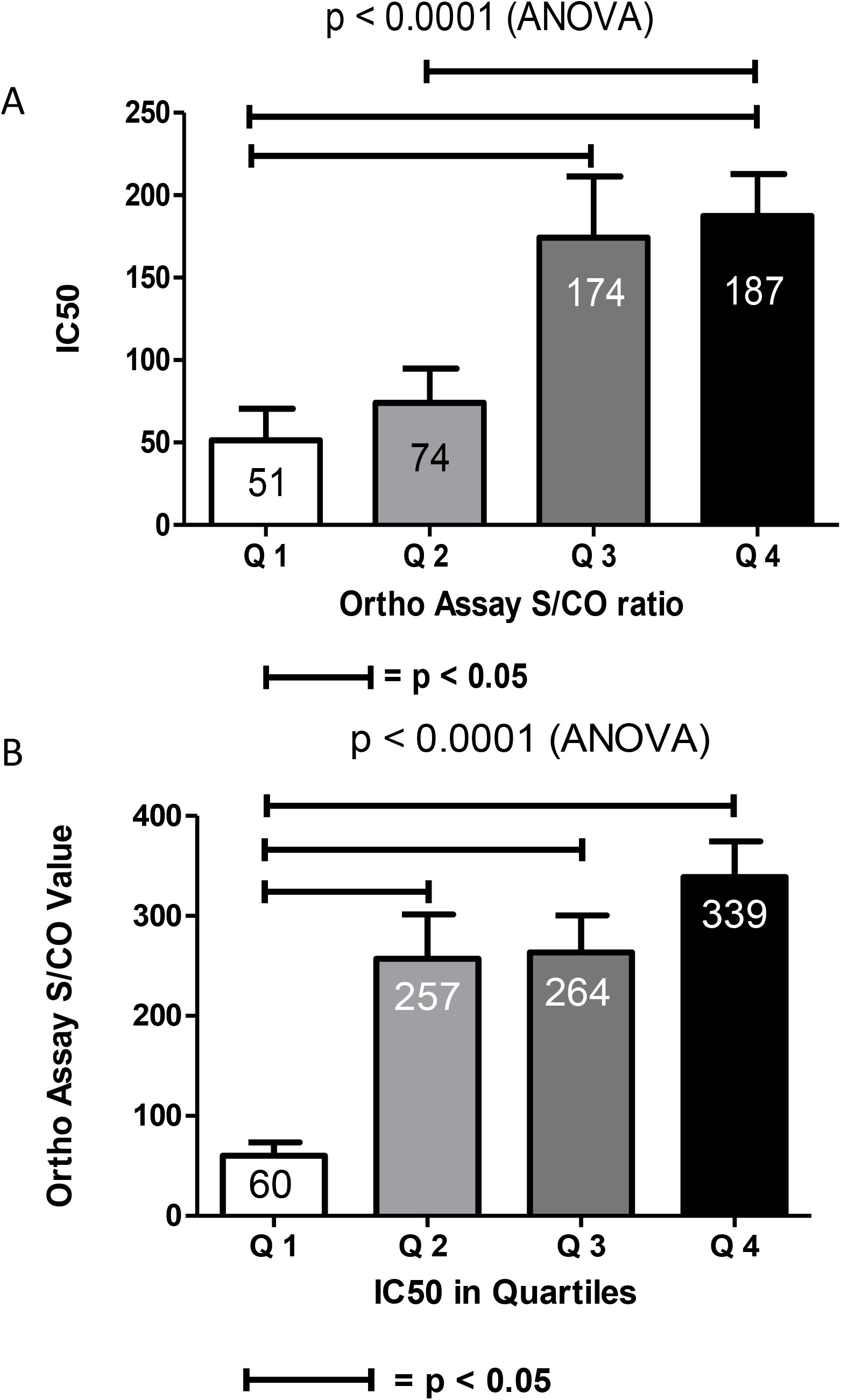
Relationship of neutralization IC50 and anti-spike total Ig S/CO values. The samples were grouped into quartiles based on S/CO values (A) or neutralization IC50 values (B) and the mean values of IC50 (A) or S/CO values (B) within each group was compared among the quartiles. The mean value within each quartile is indicated as a number within each bar. *p*-values were determined using one-way ANOVA and individual groups within the dataset were further compared using Dunnett’s test. Horizontal bars between groups indicates significance at P ≤ 0.05.

The UW Cancer Center’s biobank was able to provide plasma and/or serum samples from before and after transfusion of CCP in 19 cases. In 11 of the 19 cases (58%) there was measurable viral neutralization activity present before the transfusion of CCP. In the 8 cases (42%) that showed no measurable neutralization activity beyond the first dilution of the assay prior to transfusion, five patients (62.5%) had increased neutralization activity within 3 days of the transfusion suggestive of passive transfer of neutralization due to CCP transfusion (figure 3).

**Figure 3.**
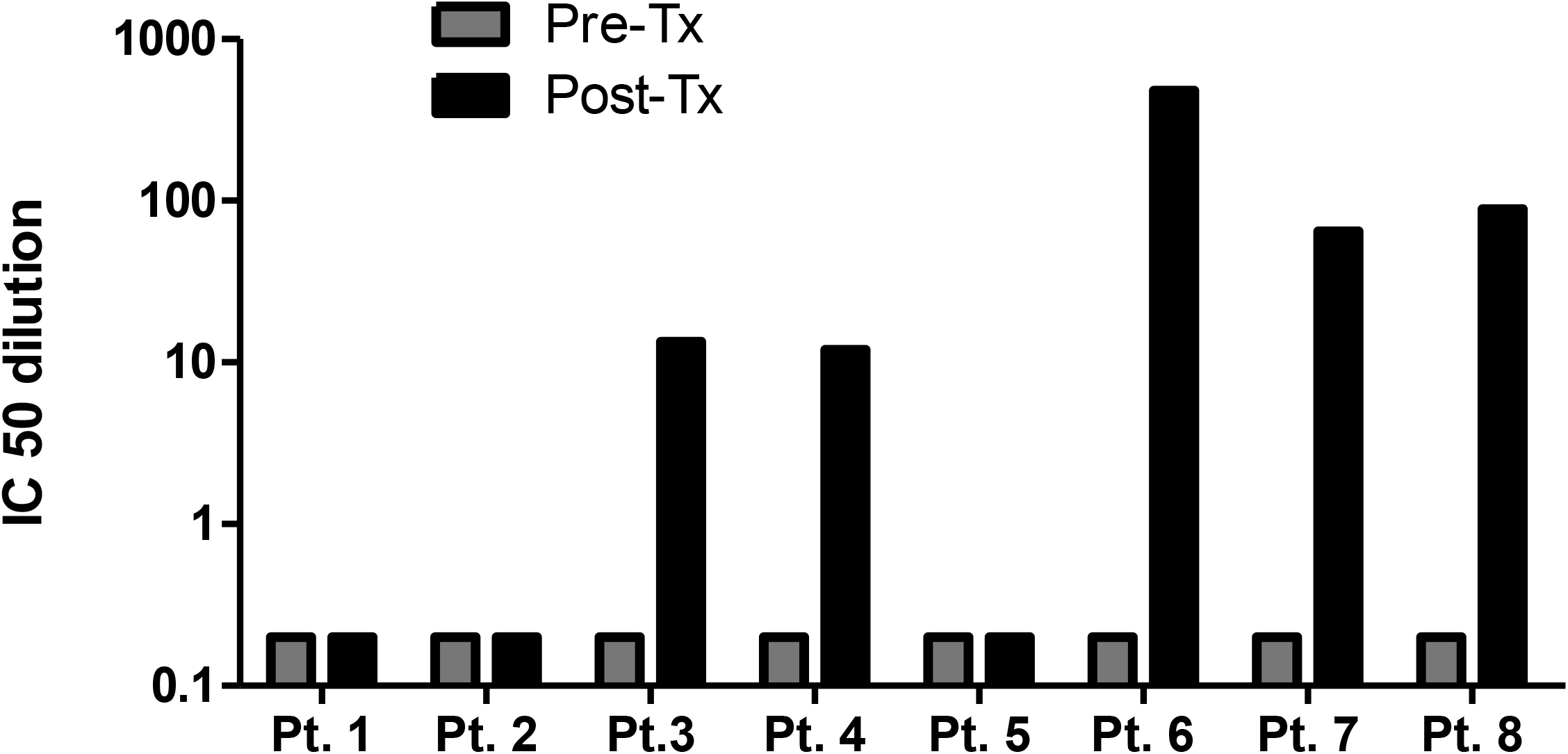
Neutralization IC50 in patient samples collected longitudinally pre- or post-plasma transfusion. Pre-transfusion (within 48 hours prior to CCP transfusion) and post-transfusion (within 3 days of CCP transfusion) samples were collected from CCP recipients. Blinded neutralization assays were performed with each sample and IC50 values were determined in each case. In the 8 cases that had no measurable neutralization activity prior to CCP transfusion, 5 of them (pts 3, 4, 6, 7 and 8) had increased neutralization activity after transfusion. (Note that when the lowest dilution of the sample did not inhibit infectivity below 50%, an arbitrary value of 0.2 was assigned to the sample to allow graphic visualization only.)

## Discussion

In COVID-19 patients, the antibody responses to SARS-CoV2 infection are variable and we confirm this with data from CCP donors and hospitalized COVID-19 patents transfused with CCP.

This variability in immune response, particularly the neutralizing antibody responses, makes selection of optimal CCP donors a challenge. Although neutralization of viral infection remains the gold standard to identify units of maximal effectiveness, this is not realistic due to the labor-and resource-intensive nature to determine neutralization activity. Until efficient high throughput assays for neutralization are available, alternative strategies to screen donors for CCP are needed. Our data support the use of commercially available serologic testing for anti-spike protein antibodies as an acceptable surrogate marker for neutralization activity to identify units for transfusion that are more likely to provide clinical efficacy. We also confirm that some patients already show neutralizing antibody responses during acute disease, questioning the role of passive immunization in these csaes. Lastly, we provide evidence of passive transfer of neutralization activity after CCP transfusion in 60% of cases without measurable neutralization before transfusion.

Studies of immune response to SARS-CoV2 infection demonstrate that some clinical characteristics are more often associated with robust antibody production and neutralization activity. The most consistent of these factors include male gender, the presence of persistent fever in the course of infections, and the severity of clinical disease. Ko et al. demonstrated that 100% of patients admitted to the hospital with COVID associated pneumonia, indicating severe disease, demonstrated higher titer neutralizing antibodies. Although neutralizing antibody titer was reduced in 94% patients presenting mild yet symptomatic disease only 80% of asymptomatic patients showed neutralizing responses ^9^. These factors may need to be considered in the selection of a CCP donor pool as many donors to date have recovered from mild or asymptomatic disease.

Whereas Wang et al. demonstrated the presence of neutralizing antibodies within 7 to 10 days of symptoms,the peak neutralization activity was not present until 33 days after infection ^13^. This study also noted that the presence of neutralizing antibodies correlated well with the presence of elevated inflammatory markers linking optimal neutralizing antibody responses to severity of disease. Lastly, this study demonstrated that antibody levels decrease by 38 % over 3 months post infection ^13^. Similar timing of antibody responses and the decline of antibody levels over time is supported in multiple studies despite considerable variability in how the response was defined ^9,14,15^. In clinical arenas the durability of the neutralization response has implications for patient recovery, the possibility of re-infection, the prospects of effective vaccines, and the efficacy of CCP.

Given the large amount of data demonstrating that many patients that do not mount a significant neutralizing antibody response after mild or asymptomatic disease, Li et al. suggested that CCP donors should be restricted to those that are greater than 28 days from the onset of symptoms and that all donors should have had disease including fever of greater than 38.5°C for more than three days^16^. With the documented decline in antibody titers over time even seriously ill patients may not have high levels of neutralizing antibodies by the time they have recovered enough to become suitable plasma donors and this may still result in sub-optimal antibody titers in the CCP donor pool.

Similar to the overall low neutralization IC50 values in our study’s donor population, Robbiani et al demonstrated that only 33% of recovered patients mounted a neutralizing antibody response with an IC50 titer of greater than 50 and that 80% of them had titers less than 1000. In this study only 1% of patients demonstrated a neutralizing antibody response with an IC50 of greater than 5000, a level we have only demonstrated in several of our acutely hospitalized CVOID-19 patients but not in any of 87 our recovered CCP donors ^17^. This study again underscores the idea that many patients may not qualify as acceptable CCP donors solely based on the fact that they had been infected and that some degree of neutralizing antibody titer assessment is needed.

A multi-center group study lead by Salazar from Houston Methodist Hospital verified that hospitalized patients with more severe COVID-19 disease created more robust antibody responses and that based on their testing system over 35% of potential convalescent plasma donors did not achieve the acceptable antibody cut-off necessary to verify viral neutralization ^18^. Based on analysis of over 250 plasma donations we come to the same conclusion that between 25% and 50% of CCP units collected locally may not have antibody levels high enough to provide therapeutic benefit. Since a plasma unit of approximately 250ml is transfused into patients with a plasma volume of approximately 2.5L, neutralizing activity of the donor unit will be expected to reduce at least 10-fold in the recipient. In this scenario, given the low antibody titers and low neutralizing activity of many units transfused early on in the national CCP programs, the therapeutic effect of high antibody titer CCP units has been underestimated in some studies.

Surrogate immune markers for viral neutralization, such as the commercially available serologic tests for anti-COVID antibodies, may narrow the acceptable donor pool. Our data reiterate multiple studies that clinically available COVID-19 serology tests modestly correlate with viral neutralization activity (Spearman’s r-value 0.29 through 0.47) ^19-22^. In a retrospective study of over 35,000 CCP transfusions in the Mayo Clinic’s EAP a sub-set analysis of 3082 cases showed that higher levels of anti-SARS-CoV2 spike IgG antibodies as measured by the Ortho IgG specific system was associated with reduced mortality. Patients receiving transfusion of units with IgG S/CO ratios above the 80^th^ percentile were more likely to survive their disease than those transfused with units containing IgG levels at or below the 20^th^ percentile of all values tested ^4^. Our current study is consistent with this finding in that we demonstrate that patients who died of their disease during their initial hospitalization were transfused CCP units with significantly lower anti-spike immunoglobulins based on the Ortho total Ig assay and that this was similar to the differences in neutralization IC50s between the groups. Joyner et al. also demonstrated that survival was improved when convalescent plasma was administered within the first 3 days of hospitalization. Our cohort supports this concept with a trend toward reduced morbidity with early transfusion.

Although our clinical patient population is small owing to the lack of a large COVID surge in our area ^2^, our study aligns with other available literature on donor qualifications for CCP. This study could be further bolstered with clinical data on the donor population to relate disease severity with immune parameters in the donor pool. However, the correlation of clinically available serology results with neutralizing titers is valuable information to select effecting CCP units. Although the study group is limited in size and lacks a control treatment group, as is the case nationwide, our observations demonstrate, for the first time, the passive transfer of neutralizing activity as was seen in 60% of our transfused patients that had no measurable neutralization activity before CCP transfusion.

Based on our results we conclude that neutralizing antibody activity can be inferred from anti-Spike antibody levels as determined by the Ortho Diagnostics systems, by either total Ig, (as we describe here), or IgG specific testing (as demonstrated by Joyner et al.) and that these values could be used to qualify CCP units that are likely to improve treatment outcomes. In addition, further evaluation of the role of anti-nucleocapsid antibodies present in CCP units and defined by the Abbott INDEX needs to be explored as our data raises questions as to the clinical/therapeutic value of this group of anti-SARS-CoV2 antibodies. Lastly, we show that just over half (58%) of the CCP recipients already had measurable neutralization activity prior to transfusion, indicating that the pre-transfusion immune response status needs to be studied as a confounding factor in understanding the clinical efficacy of convalescent plasma therapy.

## Data Availability

de-identified Data can be made available via the corresponding author

## Acknowledgment

This study was funded by Research and Development funds provided to Joseph Connor, MD by the University of Wisconsin Department of Pathology and Laboratory Medicine. The authors would like to thank the University of Wisconsin Carbone Cancer Center (UWCCC) for use of its Shared Services to complete this research. This work is supported in part by NIH/NCI P30 CA014520-UW Comprehensive Cancer Center Support. The ability of the Biobank to mobilize these COVID-19 research effort is based on leveraging established relationships with outstanding colleagues and departments including the Department of Pathology & Laboratory Medicine, the Institutional Review Board (IRB), the Institute of Clinical and Translational Research (ICTR) and the UW school of Medicine and Public Health (SMPH).

## Authorship and Conflict of Interest statements

Sanath Kumar Janaka, PhD. Study Design, assay development, experimentation, manuscript writing and editing. No conflicts of interest to report.

William Hartman, MD, PhD. Manuscript review, clinical care of subjects. No conflicts of interest to report.

Huihui Mou, PhD. Study design, assay development. No conflicts of interest to report.

Michael Farzan, PhD. Study design, assay development. No conflicts of interest to report.

Susan Stramer, MD. Study design, donor sample procurement, manuscript review and editing. No conflicts of interest to report.

Erin Goodhue, MD. Manuscript review and editing.

John Weiss, MD, PhD. Donor sample procurement/care, manuscript review and editing. No conflicts of interest to report.

David Evans, PhD Study design, assay development, manuscript review. No conflicts of interest to report.

Joseph P. Connor, MD. Study concept development, design, and coordination, regulatory /approvals, secured departmental funding, clinical data collection and analysis, statistical analysis, manuscript writing and editing. No conflicts of interest to report.

